# The presenilin 1 mutation P436S causes familial Alzheimer’s disease with elevated Aβ43 and atypical clinical manifestations

**DOI:** 10.1101/2023.12.06.23299395

**Authors:** Charles Arber, Christopher R.S. Belder, Filip Tomczuk, Rebecca Gabriele, Yazead Buhidma, Antoinette O’Connor, Helen Rice, Tammaryn Lashley, Nick C. Fox, Natalie S. Ryan, Selina Wray

**Affiliations:** Department of Neurodegenerative Disease, UCL Queen Square Institute of Neurology, London, UK; Dementia Research Centre, UCL Queen Square Institute of Neurology, London, UK; UK Dementia Research Institute at UCL, London, UK; Adelaide Medical School, The University of Adelaide, Adelaide, SA, Australia; Department of Genetics, Institute of Psychiatry and Neurology, Warsaw, Poland

**Keywords:** Familial Alzheimer’s disease, AD, PSEN1, Aβ, iPSC, PALP

## Abstract

**BACKGROUND:** Familial Alzheimer’s disease (fAD) is heterogeneous in terms of age at onset and clinical presentation. A greater understanding of the pathogenicity of fAD variants and how these contribute to heterogeneity will enhance our understanding of the mechanisms of Alzheimer’s disease more widely. The pathogenicity of the P436S mutation in *PSEN1* remains unclassified.

**METHODS:** To determine the pathogenicity of the *PSEN1* P436S mutation, we studied an expanded kindred of 8 affected individuals, with MRI (2 individuals), patient-derived iPSC models (2 donors) and post-mortem histology (1 donor).

**RESULTS:** An autosomal dominant pattern of inheritance of fAD was seen with an average age at symptom onset of 46 years and atypical features including late spastic paraparesis and non-memory led cognitive impairment in some individuals. iPSC models and post- mortem tissue supported high production of Aβ43. PSEN1 peptide maturation was unimpaired, unlike previously reported atypical mutations R278I and E280G. CONCLUSIONS: We confirm that the P436S mutation in *PSEN1* causes atypical fAD. The location of the mutation in the critical PSEN1 PALP motif may explain the early age of onset despite appropriate protein maturation.

## 1. Background

Familial Alzheimer’s disease (fAD) is caused by autosomal dominantly inherited mutations in the amyloid precursor protein (*APP*) and presenilin 1 or 2 (*PSEN1/2*) genes ^1–3^. The normal cleavage of APP by PSEN1/2 (the catalytic subunit of γ-secretase) is disrupted by fAD- associated mutations, leading to changes in amyloid-beta (Aβ) production, and predisposing the deposition of Aβ into amyloid plaques. Plaques composed of Aβ are characteristic of AD and Aβ is thought to initiate a disease cascade ^4^.

Biochemically, fAD mutations can either lead to 1) increased overall production of Aβ, or 2) elevated production of longer, more aggregation prone species such as Aβ42 and Aβ43 in relation to shorter species such as Aβ40 and Aβ38 ^5^. Aβ has been shown to be cleaved by γ- secretase in two major cleavage pathways: Aβ49>46>43>40 and Aβ48>45>42>38 ^6,7^.

Mutations in *PSEN1* destabilise the Aβ to γ-secretase complex, thereby releasing longer forms of Aβ prior to the final processing step ^8^.

More than 300 mutations in *PSEN1* have been described (alzforum.org). Mutations are distributed along the entire PSEN1 peptide and commonly cause single amino acid substitutions. Different mutations display remarkable heterogeneity in 1) age at symptom onset, which can range from 24 to over 65 years of age ^9,10^, and 2) clinical presentation: most mutations present with cognitive decline while others may present with additional motor features, such as spastic paraparesis, which can precede cognitive decline ^9,11^.

Induced pluripotent stem cells (iPSCs) offer a unique opportunity to study patient-derived neurons expressing physiological doses of mutant proteins in the cell type affected by disease ^12^. Our previous work has shown that different mutations in *PSEN1* can have distinct effects on APP processing and Aβ production; with PSEN1 protein stability and maturity affected by different mutations ^13^. Importantly, alterations in Aβ processing are conserved between iPSC models and patient plasma ^14^, validating the relevance of iPSCs to model Aβ production in patients.

Aβ43 is a particularly aggregation-prone species ^15^ that is commonly overlooked. Employing iPSC models, work by us and by others has demonstrated that mutations in *PSEN1* such as R278I, E280G and L435F lead to an increased relative production of Aβ43 compared to shorter peptides ^13,16–18^. These mutations frequently have atypical clinical presentations, with prominent motor features including spastic paraparesis or extrapyramidal signs ^9^.

In this study, we set out to investigate the P436S mutation in *PSEN1*. Despite two individuals with memory impairment having been previously described with this mutation ^19^, the pathogenicity of P436S remains unclassified (alzforum.org). Here, using an expanded kindred, clinical findings, post-mortem brain tissue and iPSC models, we tested the hypothesis that *PSEN1* P436S is associated with fAD, and further characterise its clinical manifestations.

## 2. Methods

### 2.1 Subjects

We provide an updated three generation pedigree of a family with *PSEN1* P436S associated fAD. Clinical information was obtained from research and clinical contact. Post-mortem results were available for one individual (individual II-4, Fig 1).

**Figure 1.**
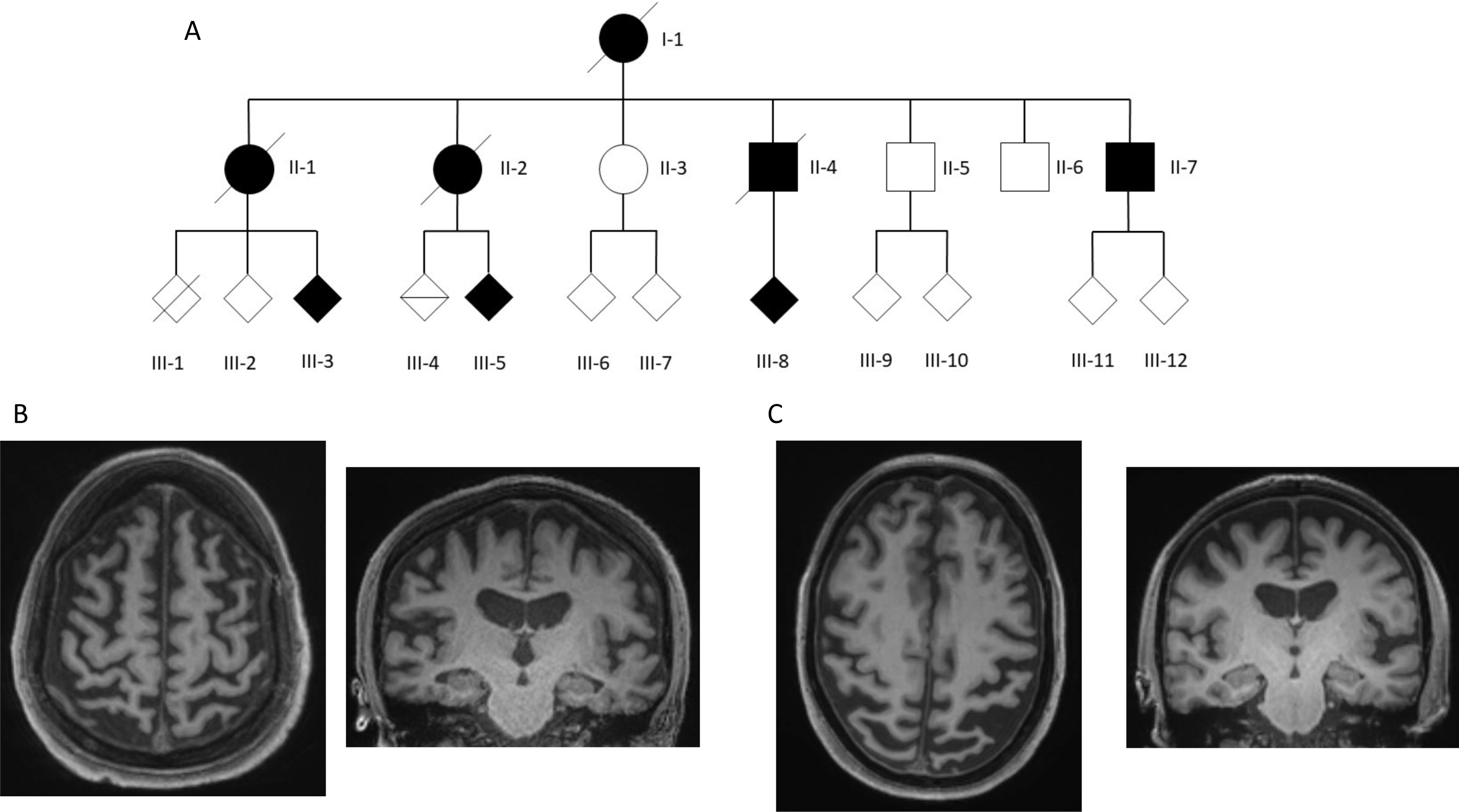
*PSEN1* P436S expanded family kindred and MRI findings support autosomal dominant fAD. **A)** Updated Kindred of family. II-4 is the individual previously reported by Palmer et al., 1999. **B)** Example MRI images from individual III-5. T1 axial and coronal images are shown at age, demonstrating generalised atrophy and low volume hippocampi. **C)** Example MRI images from individual III-8. T1 axial and coronal images are shown at age, demonstrating posterior cerebral atrophy, widening of the left sylvian fissure and relatively preserved hippocampal volume.

Fibroblast biopsies were taken with research ethics approval (11/LO/0753) and with informed consent.

### 2.1 Cell culture

Fibroblasts were grown in DMEM with 10% foetal bovine serum and passaged using 0.05% trypsin. Fibroblasts were reprogrammed using Epi5 plasmids, obtained from Thermo Fisher. A Lonza P2 nucleofector was used to electroporate the plasmids into 750,000 fibroblasts.

Media was changed from fibroblast media to Essential 8 iPSC media after passaging the fibroblasts onto Geltrex substrate at day 7. Colonies emerged after 4-5 weeks and were picked, expanded and banked for characterisation. iPSCs were routinely cultured in Essential 8 media on Geltrex substrate and passaged using 0.5mM EDTA at a ratio or around 1:6 every 5 days.

Neuronal differentiation was performed as previously reported ^13,20,21^. Briefly, iPSCs were grown to confluence and then switched to N2B27 media with 10μM SB431542 and 1μM dorsomorphin. After 10 days of neural induction, neural precursors were passaged onto laminin in N2B27 media. A final passage of progenitors was performed at day 35 onto laminin and poly-ornithine coated wells. The cells were allowed to terminally differentiate and mature until day 100, which was taken as the final time point.

All reagents were Thermo Fisher unless stated otherwise.

### 2.2 Sanger sequencing

Sanger sequencing was performed with SourceBio on PCR amplicons (GoTaq, Promega) of genomic DNA using the following primers: forward primer GTCTTTCCCATCTTCTCCAC and reverse primer GGGATTCTAACCGCAAATAT.

### 2.3 qPCR-based karyotyping

qPCR-based karyotyping was performed using the Stem Cell Technology human pluripotent stem cell genetic analysis kit. This tests for the 8 most common chromosomal amplifications/deletions in stem cell cultures.

### 2.4 qPCR

RNA was extracted from neurons using Trizol and RNA precipitation. 2μg of RNA was reverse transcribed to cDNA using Superscript IV following manufacturer’s instructions. qPCR was performed using POWER Sybr green using the following primers all with annealing temperatures at 60°C: housekeeping gene *RPL18A* forward CCCACAACATGTACCGGGAA and *RPL18A* reverse TCTTGGAGTCGTGGAACTGC; *TBR1* forward AGCAGCAAGATCAAAAGTGAGC and *TBR1* reverse ATCCACAGACCCCCTCACTAG; *CTIP2* forward CTCCGAGCTCAGGAAAGTGTC and *CTIP2* reverse TCATCTTTACCTGCAATGTTCTCC; *TUBB3* forward CATGGACAGTGTCCGCTCAG and *TUBB3* reverse CAGGCAGTCGCAGTTTTCAC; *APP* forward GGTACCCACTGATGGTAAT and *APP* reverse GGTAGACTTCTTGGCAATAC; *PSEN1* forward TATCAAGTACCTCCCTGAAT and *PSEN1* reverse ACCATTGTTGAGGAGTAAAT; *PSEN2* forward GACTCCTATGACAGTTTTGG and *PSEN2* reverse GCACACTGTAGAAGATGAAGT.

### 2.5 Immunocytochemistry

Cells were plated on glass coverslips and fixed using 4% paraformaldehyde for 15 minutes at room temperature. Cells were then permeabilised in PBS with 0.3% Triton-X-100 (hereafter PBSTx) via three washes. Cells were then blocked in 3% BSA in PBSTx for 30 minutes at room temperature. Primary antibodies were added in blocking solution overnight at 4°C. Cells were then washed three times in PBSTx and secondary antibodies (AlexaFluor) were added in blocking solution for 1 hour at room temperature. Cells were washed a final three times in PBSTx and counterstained using DAPI. Finally, cells were mounted, using DAKO fluorescence mounting media and imaged on a Zeiss LSM microscope with no post-hoc adjustments to the images.

Primary antibodies were: SSEA4 Biolegend 330401 RRID:AB_1089209; NANOG Cell Signalling Technology 4903 RRID:AB_10559205; TBR1 Abcam ab31940 RRID:AB_2200219; TUJ1 Biolegend 801201 RRID:AB_2313773).

### 2.6 Immunohistochemistry

Post-mortem brain tissue from individual II-4 assessed in this study was obtained through the brain donation programme of the London Neurodegenerative Diseases Brain Bank. The protocols used for brain donation and ethical approval for this study were approved by a London Research Ethics Committee and tissue is stored for research under a license from the Human Tissue Authority. The standard diagnostic criteria for the neuropathological diagnosis of Alzheimer’s disease were used ^22–24^. Slides with 8µm paraffin embedded tissue sections were incubated at 60°C overnight. Sections were deparaffinised in Xylene and rehydrated in decreasing grades of alcohol. Slides were incubated in methanol/hydrogen peroxide (0.3%) solution for 10min to block endogenous peroxidase activity. Slides were then incubated in 98% formic acid for 15min. For heat-induced antigen retrieval, slides were then transferred to a boiling solution of 0.1M citrate buffer (pH6.0) and pressure cooked at maximum pressure for 10min. Subsequently sections were incubated in 1% bovine serum albumin for 30min at room temperature to block non-specific binding. Sections were incubated with primary antibody for 2h at room temperature. The antibodies used for IHC in this study were mouse-derived monoclonal anti-β-amyloid, 1-43 and anti-β-amyloid, clone 6F/3D (BioLegend, 805607, 1:500 and DAKO, M0872, 1:200). After three 5min washes in tris-buffered saline with 0.1% Tween-20 (TBS-Tw); slides were incubated for 1 h in biotinylated goat anti-mouse IgG secondary antibody (Vector Laboratories BA 9200, 1:200). Slides were washed as before and then incubated in pre-conjugated Strept(avidin)–Biotin Complex (ABC; DAKO) for signal amplification. The slides were then washed for a final time before being submerged in 3,3ʹ-Diaminobenzidine (DAB) chromogen and then counterstained in Mayer’s haematoxylin (BDH). Finally, slides were dehydrated in increasing grades of alcohol (70, 90 and 100% IMS), cleared in xylene and mounted. Tissue sections were digitally scanned using an Olympus VS120 slide scanner at 20 × magnification.

### 2.7 Western blotting

Cells were lysed in RIPA buffer containing protease and phosphatase inhibitors (Roche). Lysates were centrifuged to remove insoluble debris and protein content was quantified using BCA assay (BioRad). Samples were standardised for protein content and denatured using LDS buffer and dithiothreitol with boiling at 95°C for 5 mins. Proteins were separated using 4-12% precast polyacrylamide gels and transferred onto nitrocellulose membranes. Membranes were blocked in PBS with 0.1% Tween-20 (PBSTw) with 3% BSA. Primary antibodies were added overnight at 4°C in blocking solution. Membranes were then washed three times and secondary antibodies (Licor) were added for 1 hour in blocking solution. A final three washes were performed prior to imaging on a Licor Odyssey scanner.

Primary antibodies were: PSEN1 NTF Millipore MAB1563 RRID: B_11215630; sAPPβ IBL JP18957 RRID:AB_1630824; sAPP total (22c11) Millipore MAB348 RRID:AB_94882; APP ctf Thermo A8717 RRID:AB_258409; PSEN2 Cell Signalling Technology 9979 RRID:AB_10829910; Actin Sigma A1978 RRID:AB_476692.

### 2.8 Aβ ELISAs

Media was harvested after 48 hours conditioning on iPSC-derived neurons. Conditioned media was centrifuged at 2,000g for 5 minutes to remove any cellular debris. Aβ38, Aβ40 and Aβ42 was quantified simultaneously via Meso Scale Discovery V-PLEX Aβ peptide panel (6E10). Samples were diluted 2-fold and measurements were made against standard curves on an MSD Sector 6000. Aβ43 was quantified using IBL Amyloid Beta (1-43) (FL) ELISA. Media was run undiluted against a standard curve following manufacturer’s protocols.

Measurements were made on a TECAN SPARK 10M plate reader.

## 3. Results

### 3.1 Expanded kindred and case descriptions

We provide an updated 3 generation pedigree of a family with *PSEN1* P436S (Fig 1A). Clinical information was obtained from research and clinical contact (Table 2).

The kindred are consistent with an autosomal dominant pattern of inheritance (Fig 1A). The average age of onset is 46 years (44-50), most commonly with a memory led decline, and with apraxia as a common associated feature. Leading executive dysfunction was observed in one individual. Lower limb pyramidal signs or spastic paraparesis as a late manifestation was observed frequently; in 3 of the 5 affected individuals with documented neurological examinations. MRI was available for two individuals (Fig 1B-C) and demonstrates generalised cerebral atrophy including hippocampi for individual III-5 and marked posterior atrophy with relatively preserved hippocampal volume for individual III-8. Neither had white matter hyperintensities on fluid attenuated inversion recovery (FLAIR) or cerebral microbleeds on susceptibility weighted imaging (SWI). Together, these findings support the hypothesis that *PSEN1* P436S mutations cause fAD, with observed atypical features in some individuals including late spastic paraparesis and one individual with non-episodic memory lead cognitive decline in combination with significant posterior atrophy on imaging.

### 3.1 iPSC models demonstrate elevated Aβ43:40 ratios in P436S mutant neurons

In order to generate a human neuronal model of the P436S mutation, fibroblasts from two donors were reprogrammed to iPSCs (III-5 and III-8, see Table 1). Newly generated iPSCs were confirmed to have iPSC morphology and expression of pluripotency-associated factors NANOG and SSEA4 by immunocytochemistry (Fig 2A). The presence of the P436S mutation was confirmed by Sanger sequencing and the iPSC lines were shown to have a stable karyotype (Fig S1A-B). Together with two control iPSC lines, cells were differentiated to cortical, glutamatergic neurons. Patient-derived lines generated iPSC-derived neurons with a similar efficiency to control lines, suggesting that the mutation did not affect neuronal differentiation (Fig 2A and Fig S1C-E).

**Figure 2.**
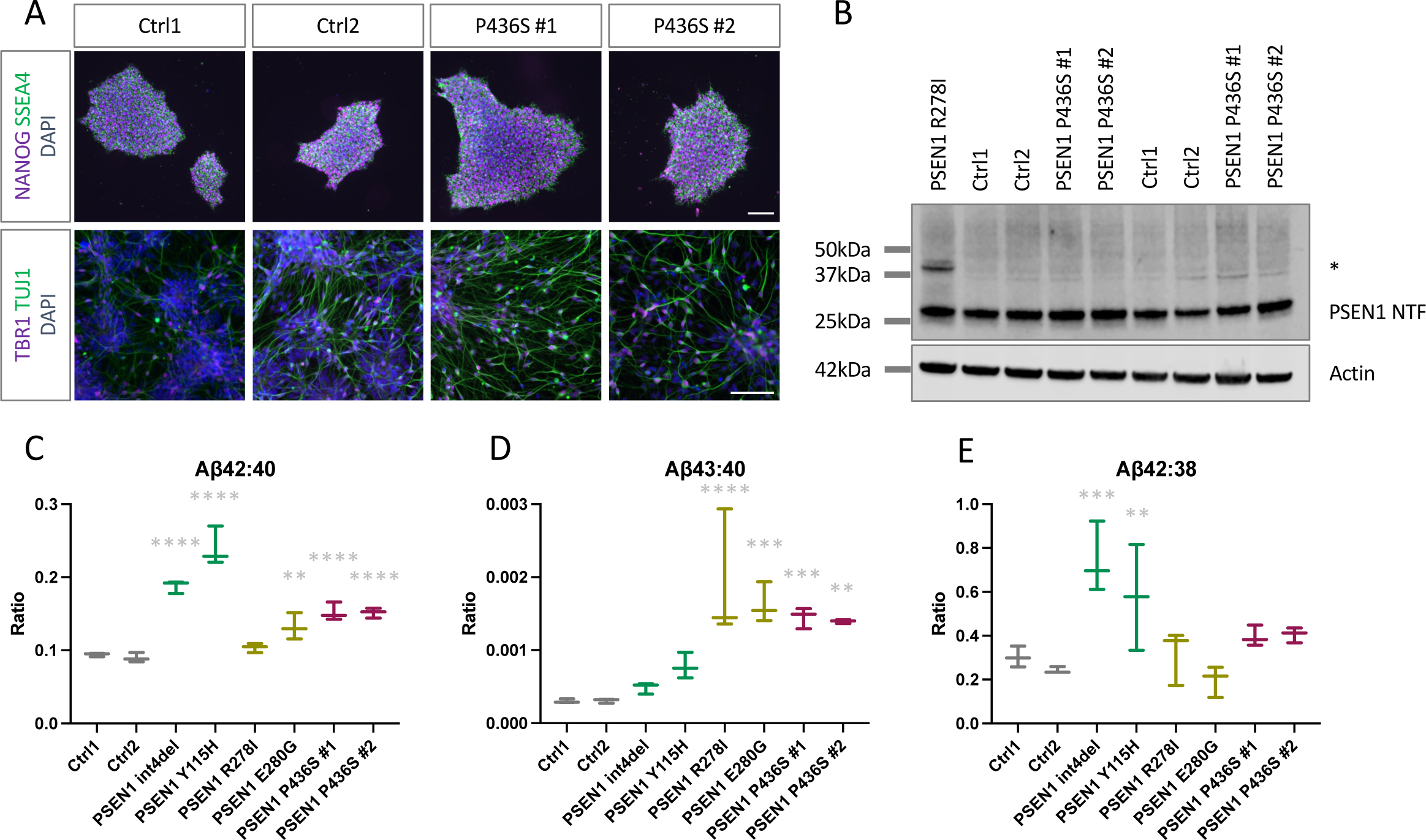
*PSEN1* P436S patient-derived iPSC neuron models display fAD-associated Aβ profiles **A)** Immunocytochemical analysis of iPSCs (top) confirming expression of pluripotency markers NANOG and SSEA4. Immunocytochemical analysis of iPSC-derived neurons at day 50 of differentiation, bottom, displaying neuronal morphology, expression of deep layer cortical marker TBR1, and neuronal-specific tubulin TUJ1. **B)** Western blot analysis of PSEN1 maturation neuronal lysates collected after 100 days of differentiation. Immature PSEN1 at 48kDa (*) is cleaved to a mature N-terminal fragment (NTF) at around 28kDa. *PSEN1* R278I is displayed as a mutation that causes maturation defects. Actin is used as a loading control. Two (of three) independent inductions are shown. **C-E)** Aβ peptide analysis by electrochemiluminescence to depict the disease-associated Aβ42:40 ratio, the Aβ43:40 ratio and the processivity ratio Aβ42:38. Int4del and Y115H (green) display typical *PSEN1* mutation-associated ratio changes, R278I and E280G (yellow) are shown as high Aβ43 producing mutations; each serving as comparators to control (grey) and P436S (red) lines. P436S lines display significantly raised Aβ42:40 and Aβ43:40 ratios. Data represents three independent inductions for each line. Significance represents ANOVA and Dunnett’s multiple comparisons, ** p < 0.01, *** p < 0.001, **** p < 0.0001.

**Table 1.** Fibroblast/iPSC donor information.

|  | Gender | Kindred member | Age at onset | Age at biopsy | Status |
| --- | --- | --- | --- | --- | --- |
| P436S.1 | Female | III-5 |  |  | Symptomatic |
| P436S.2 | Female | III-8 |  |  | Symptomatic |

**Table 2.** Case descriptions of the expanded *PSEN1* P436S kindred.

| Individual | Age at onset | Age at death | Clinical observations |
| --- | --- | --- | --- |
| I-1 | Unknown | ✖ | Diagnosed with “pre-senile dementia”. |
| II-1 | ✖ | ✖ | Initial memory led decline and apraxia. On examination age ✖, pathologic hyperreflexia in the lower limbs and diffuse cortical atrophy noted on CT scan. |
| II-2 | ✖ | ✖ | Initial memory impairment. MMSE at age ✖ was 22/30, with impaired recall, calculation, sentence and figure copying. There was late development of hallucinations and seizures. |
| II-4 | ✖ | ✖ | Initial memory impairment. MMSE age ✖ was 7/30, with severe verbal, visual and general memory and attention impairment on psychometry. CT brain demonstrated moderate generalized atrophy. Post-mortem confirmation of AD neuro-pathologic change (see Fig 3). Confirmed carrier of *PSEN1* P436S. |
| II-7 | ✖ | N/A | Limited information. |
| III-3 | ✖ | N/A | Initial impairment in memory, with word-finding difficulties and apraxia. |
| III-5 | ✖ | N/A | Initial memory impairment. At age ✖ they developed apraxia and dyscalculia. On examination age ✖ there was apraxia, myoclonus, increased tone in the lower limbs with hyper-reflexia and downgoing plantar responses, and no parkinsonism or cerebellar signs. MRI aged ✖ demonstrated generalized atrophy including the hippocampi (Fig 1B). At age ✖ they developed word-finding difficulty and a stutter. Confirmed carrier of *PSEN1* P436S. |
| III-8 | ✖ | N/A | Initial executive dysfunction with slowed processing and planning, then developed topographic memory dysfunction, apraxia and language dysfunction with word-finding difficulties. On examination aged ✖ there<br>was apraxia, spasticity and hyper-reflexia in the lower limbs with downgoing plantar responses, subtle myoclonus. MMSE was 14/30. MRI aged [REDACTED] demonstrated posterior atrophy (Fig 1C). They developed agitation aged [REDACTED] treated with risperidone, and on examination age [REDACTED] there was parkinsonism with hypomimia, right sided rest tremor, rigidity and bradykinesia. They developed seizures age [REDACTED]. Confirmed carrier of *PSEN1* P436S. |

PSEN1 undergoes autoproteolysis from an immature full-length peptide of around 48kDa to two mature peptides of around 28kDa and 18KDa. Some mutations associated with atypical presentations of fAD have shown deficits in PSEN1 protein maturation ^13,17^. To investigate the maturation of P436S-variant PSEN1, western blotting of neuronal lysates was performed. Uncleaved, immature PSEN1 is observed at around 48kDa, whereas mature, cleaved PSEN1 is observed at around 28kDa. In contrast to PSEN1 R278I, results suggested appropriate processing of the PSEN1 P436S peptide to mature peptide fragments (Fig 2B).

Analysis of Aβ species released into the culture media by the iPSC-derived neurons demonstrated that both lines with the P436S mutation displayed a raised Aβ42:40 ratio and a large increase in the Aβ43:40 ratio (Fig 2C-D). The processivity-associated Aβ42:38 ratio was similar in P436S neurons compared with controls (Fig 2E), suggesting a predominant effect on the Aβ49>46>43>40 processing pathway. When normalised for cell numbers, all Aβ species measured showed an increased concentration in the cell culture media compared to other mutations (Fig S1F-I). This contrasts with previously studied *PSEN1* mutations which show decreased generation of shorter Aβ peptides such as Aβ38, attributable to reduced processivity (Fig S1F-I). *APP* expression, *PSEN1* expression and amyloidogenic versus non-amyloidogenic processing were not altered and therefore could explain this finding (Fig S3).

Together, these data support the finding that *PSEN1* P436S leads to impaired Aβ processing and high Aβ43 production, similar to some other fAD mutations with atypical clinical presentations.

### 3.1 Pathological diagnosis and confirmation of Aβ43 pathology in a P436S carrier

Microscopic investigation of individual II-4 confirmed the presence of severe Alzheimer’s disease-type pathology, using the National Institute on Aging/Alzheimer’s Association criteria. Microscopic investigations demonstrated frequent and widespread deposition of Aβ in parenchymal plaques (corresponding to Thal phase 5), the neuritic plaque pathology was of CERAD frequent degree and also in leptomeningeal and parenchymal blood vessels, indicative of severe cerebral amyloid angiopathy (Fig 3). The tau pathology was very severe throughout including the striate and peristriate cortices. The neurofibrillary tangle pathology, therefore, corresponded to Braak and Braak stage VI. In summary this case had ‘High’ Alzheimer’s disease neuropathological change with an NIA/AA score A3B3C3. In addition, there was also evidence of amygdala predominant Lewy body disease. There was no evidence of TDP43 pathology.

**Figure 3.**
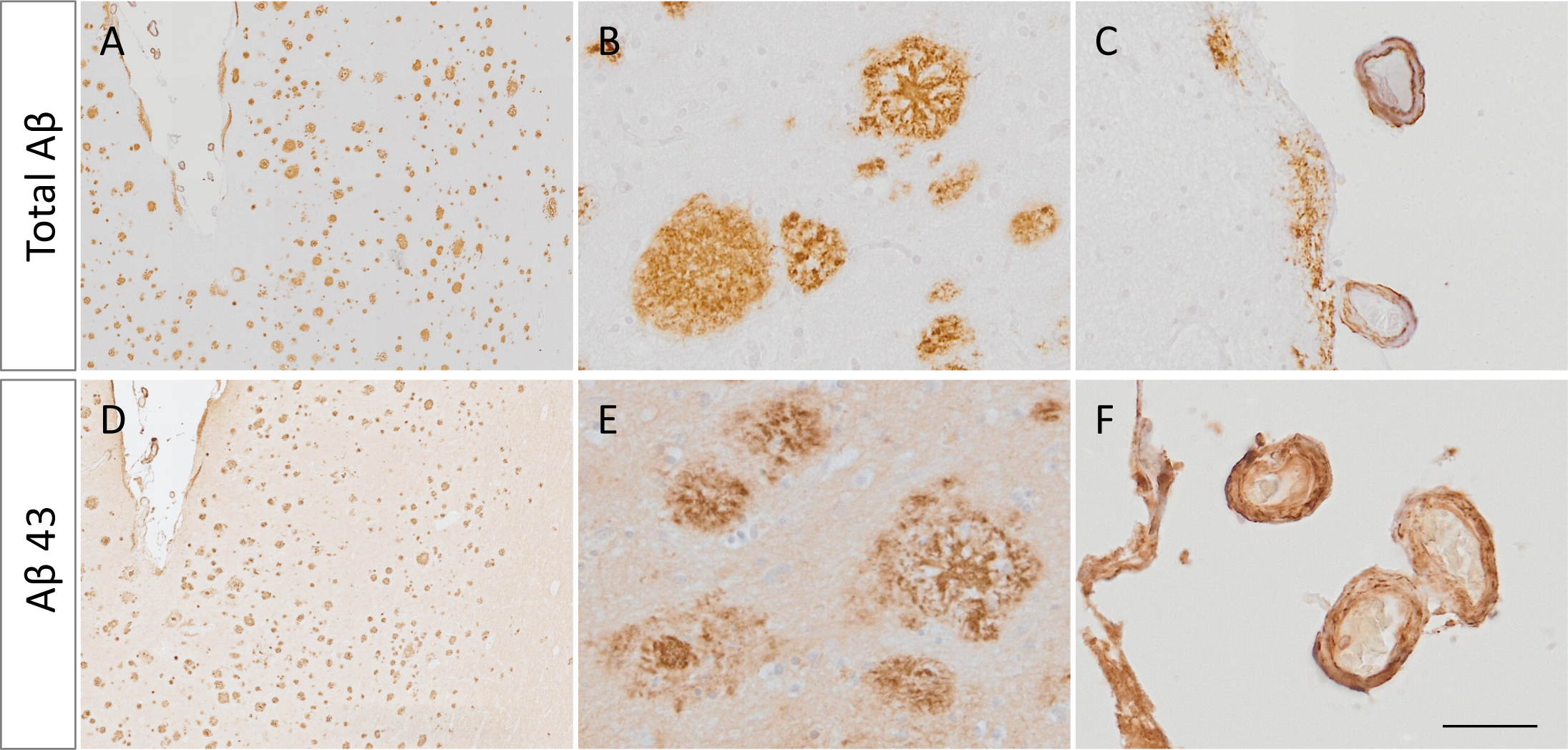
**Immunohistochemical staining of post-mortem tissue displays A**β**43 deposition. A-C)** Total Aβ staining shows plaque pathology (A-B) and amyloid angiopathy (C). **D-E)** Aβ43-specific staining demonstrates substantial Aβ43 deposition in both parenchymal plaques and in vessels as cerebral amyloid angiopathy. Scale bar represents 500µm in A and D; 50µm in B, C, E and F.

To confirm the finding that *PSEN1* P436S mutant neurons produce high levels of Aβ43, we also performed immunohistochemical analysis for total Aβ and Aβ43 on post-mortem brain tissue. Analysis suggests that Aβ43 represents a major component of the amyloid plaques in P436S-associated tissue. Aβ43 staining highlights dense cores within plaques as well as extensive cerebral amyloid angiopathy. There was also evidence for cotton wool plaque pathology (round lesions that lack central dense cores that have been linked with atypical *PSEN1* mutations), although cotton wool plaques and dense core plaques were present with similar abundance. These data confirm that Aβ43 pathology is associated with the *PSEN1* P436S mutation.

## 4. Discussion

Making use of an expanded family history, MRI brain scans, iPSC models and post-mortem tissue, we confirm that the P436S mutation in *PSEN1* is causative of familial Alzheimer’s disease and describe its atypical clinical features.

This kindred highlights the heterogeneity that can be observed in fAD despite a common causative mutation, and the potential atypical cognitive presentations and motor syndromes, particularly with certain *PSEN1* mutations ^9^. While the age at symptom onset was relatively consistent (range 44-48), the disease duration (range 5-21) and age at death (range 49-68) varied widely. This is in keeping with previous work in fAD demonstrating that survival time is influenced by mutation to a lesser extent than age at onset ^25^. In most cases the cognitive syndrome was memory led (4/5), but one individual (III-8) presented with an atypical cognitive manifestation of initial executive dysfunction (a clinical phenotype that can be seen as an atypical manifestation of fAD and sporadic early onset AD) ^9,26,27^, and demonstrated significant posterior cerebral atrophy on imaging with relative preservation of the hippocampi (radiologically similar to that which is seen in posterior cortical atrophy (PCA), another atypical clinical phenotype of early onset AD which is held to be sporadic ^28^, although notably this individual did not have the clinical syndrome of PCA). Distinct from sporadic AD and fAD caused by *APP* mutations, in this kindred spastic paraparesis was observed as a late manifestation in a majority of cases with documented neurological examinations (3/5), and this study further reinforces the need to consider *PSEN1* mutations in individuals presenting with cognitive decline and spastic paraparesis ^29^. Late parkinsonism was seen in one individual, although this could be confounded by neuroleptic exposure.

The link between Aβ43 and atypical motor manifestations warrants attention in future clinical studies. Elevated Aβ43 has been reported in other fAD mutations that are associated with spastic paraparesis, including E280G and R278I ^13,17^, and with parkinsonism, including L435F ^30,31^. Our study also supports the correlation between *PSEN1* mutations associated with spastic paraparesis and cotton wool plaque pathology, which in turn has been shown to correlate with cerebral amyloid angiopathy in some, but not all, cases ^31,32^. The pathophysiology of spastic paraparesis in *PSEN1*-associated fAD is not well understood and may relate to altered processing of γ-secretase substrates other than amyloid precursor protein ^29^. The apparent association between increased Aβ43 and spastic paraparesis may potentially represent a signature of altered γ-secretase function as it relates to these other substrates.

The PSEN1 P436S peptide variant shows appropriate autoproteolysis and maturation, shown via the absence of an immature PSEN1 band by western blotting. This replicates findings in mouse embryonic fibroblasts and cell models, whereby P436S shows largely normal processing, which is in contrast to the P436Q mutation ^31,33^. The current work disputes a previous correlation between incomplete PSEN1 peptide maturation and Aβ43 production for the R278I and E280G mutations ^13,17^.

The P436 residue of PSEN1 lies within the PALP motif (residues P433, A434, L435 and P436; reviewed by Bagaria, Bagyinszky and An, 2022). Stereologically, the PALP motif lies close to the catalytic pore of PSEN1 and is critical for proper enzymatic function ^35,36^. The importance of this region may explain the early age at onset of mutations associated with this motif; P433S leads to age at onset of 34 years ^33^, A434C and A434T have ages at onset from 29 and 35 years respectively ^37,38^, L435F has an age of onset of 47 ^39^ and P436Q has an age at onset of 28 ^40^. Therefore, we theorise that the P436S mutation may have a less severe impact on PSEN1 function compared to P436Q (which shows defective PSEN1 autoproteolysis, a very early age at onset, and is also associated with spastic paraparesis and posterior cerebral clinical features ^31,40^) yet still cause fAD with a relatively early age at onset due to the importance of the PALP motif. Proline residues provide structural rigidity and are often crucial to peptide turns. This potentially highlights 1) the importance of the PALP motif close to the ninth transmembrane domain of PSEN1, and 2) a more detrimental effect of the larger glutamine residue in P436Q compared to the smaller serine substitution in P436S. Finally, we were surprised to see increased levels of all Aβ species measured in P436S conditioned media. This finding supports data from cell models ^33^ which show increased Aβ40, Aβ42 and Aβ43 production. Further experiments are required to test the basis of this finding.

In summary, this study confirms the pathogenicity of the *PSEN1* P436S mutation, validating a causative effect for fAD. The data further underline the relevance of Aβ43 in the pathogenesis of fAD, the association between Aβ43 and atypical motor manifestations, and support a critical importance of the PALP domain in PSEN1 enzymatic function. Together, these findings further develop our molecular understanding of fAD and highlight the importance of considering clinical heterogeneity when designing clinical studies.

## Data Availability

All data produced in the present study are available upon reasonable request to the authors.

## Acknowledgements

We wish to thank the individuals and their families for contributing to this work and the London Neurodegenerative Diseases Brain Bank for access to the post mortem brain tissue. The London Neurodegenerative Diseases Brain Bank receives funding from the MRC and as part of the Brains for Dementia Research program, jointly funded by Alzheimer’s Research UK and Alzheimer’s Society.

---

**Supplementary Figure 1.**
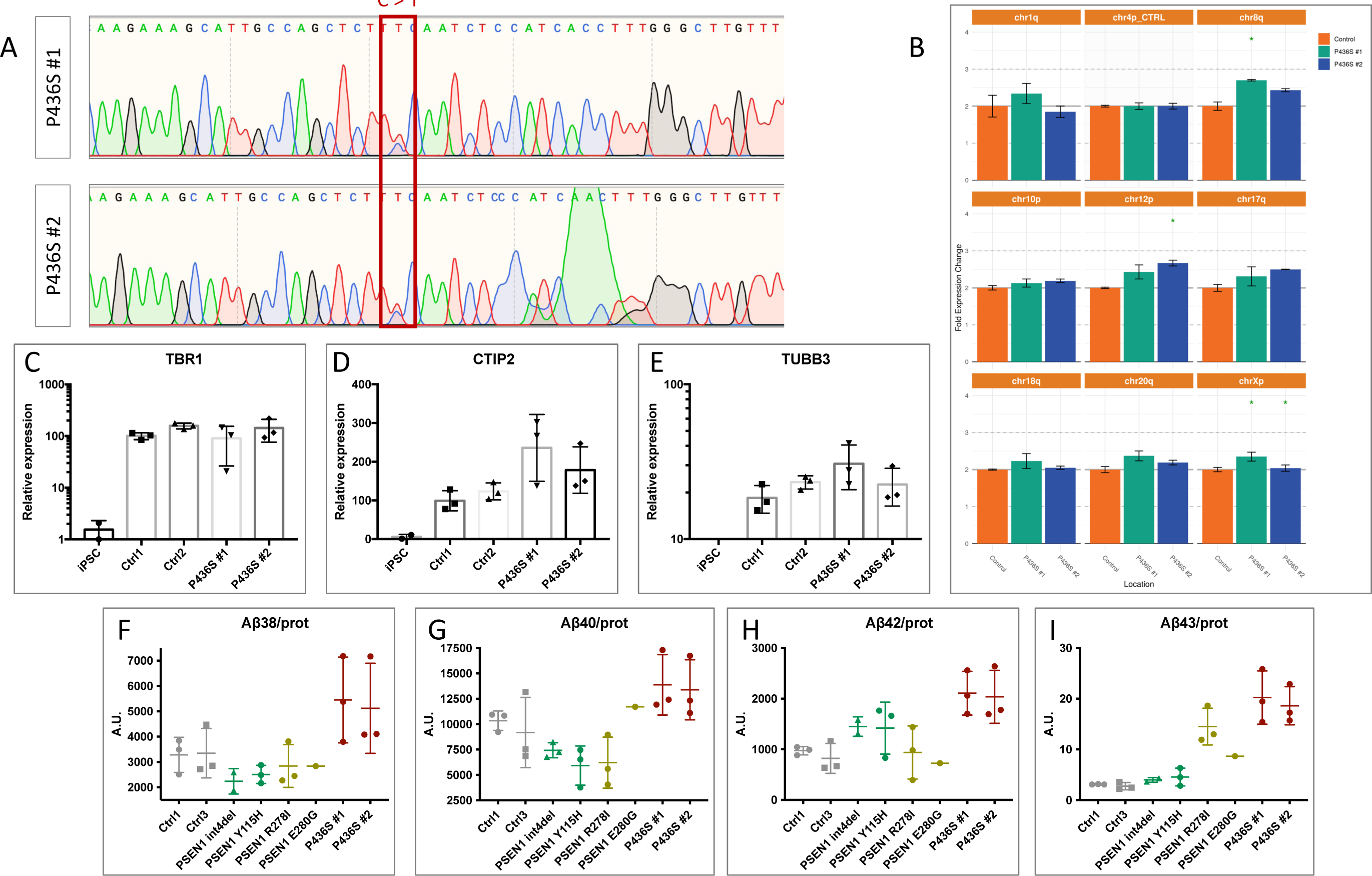
iPSC model characterisation. **A)** Sanger sequencing of *PSEN1* in iPSCs confirms the presence of the C>T mutation. **B)** PCR-based karyotype analysis suggests a normal karyotype of the two P436S lines. **C-E)** qPCR analysis of cortical marker expression in day 100 iPSC-derived neurons supports appropriate neuronal specification. TBR1, CTIP2 and TUBB3 are markers of deep layer cortical neurons, middle layer cortical neurons and pan-neuronal tubulin, respectively. **F-I)** Aβ species quantification (pg/ml) normalised to the protein content of the corresponding neurons to estimate overall Aβ production. P436S mutations produce high levels of all species.

**Supplementary Figure 2.**
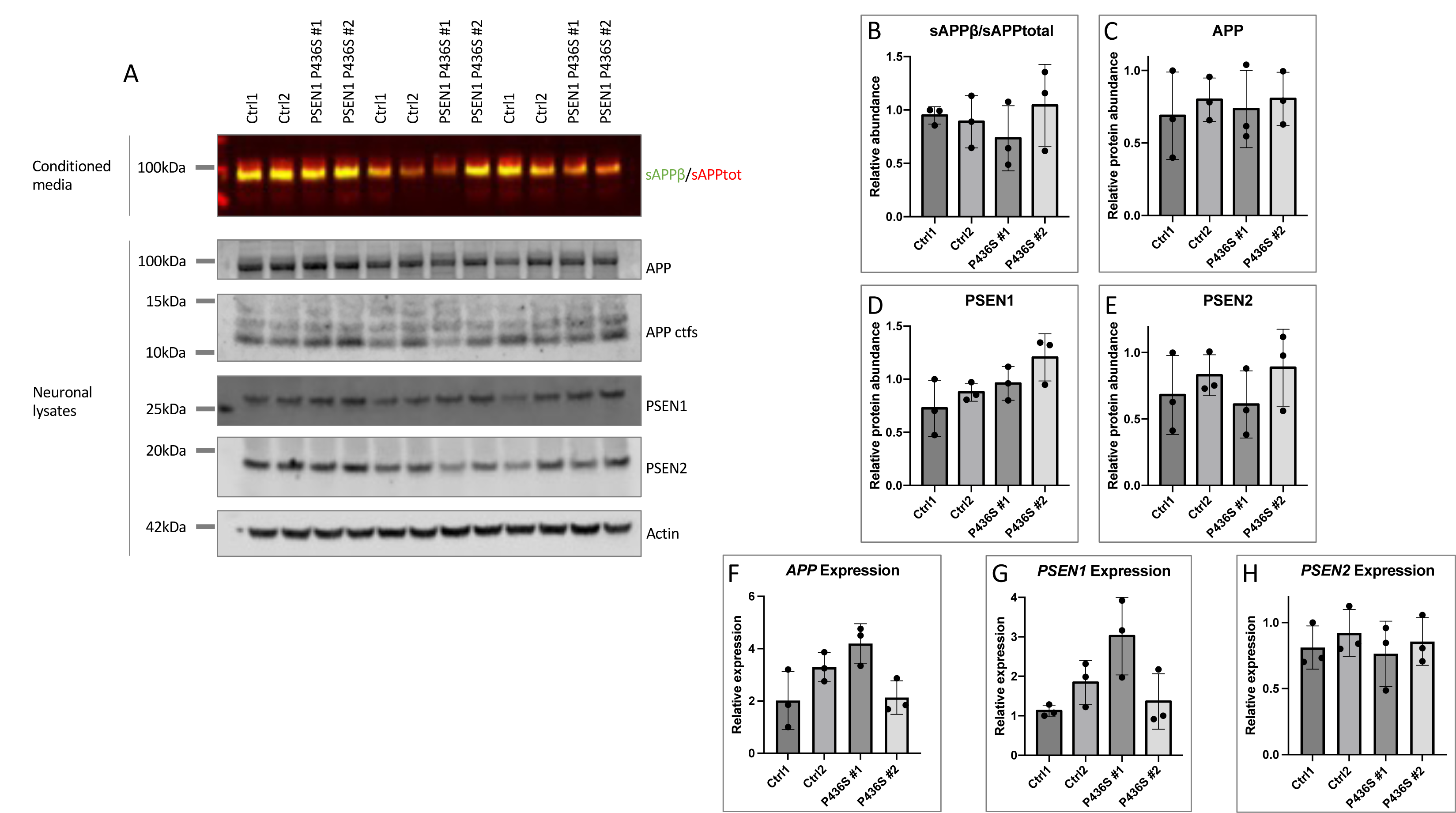
APP and PSEN1 expression in iPSC models. **A-E)** *PSEN1* P436S mutations do not affect amyloidogenic versus non-amyloidogenic processing, based on sAPPβ production levels, and do not affect APP, PSEN1 or PSEN2 protein level. **F-H)** APP, PSEN1 and PSEN2 expression levels are not significantly affected by the P436S mutation. Protein and RNA represent three independent neuronal inductions for each line.

